# STREAM-SMR: Sequential Bayesian state-space monitoring of standardized mortality ratios — a simulation comparison with risk-adjusted CUSUM and EWMA

**DOI:** 10.64898/2026.08.18.26360662

**Authors:** Kunihisa Ohno

## Abstract

**Background:** Sequential monitoring of risk-adjusted mortality in intensive care typically relies on alarm-generating control charts — the risk-adjusted CUSUM, EWMA, or VLAD. These charts signal deterioration but do not return what clinicians and registry stewards ultimately need to interpret: a calibrated, continuously updated estimate of the standardized mortality ratio (SMR) itself.

**Methods:** STREAM-SMR is a conjugate gamma–Poisson dynamic generalized linear model in which the latent log-SMR evolves through a discount factor δ and the alarm statistic is the posterior exceedance probability P(SMR > 1). The construction is closed-form, exact for zero-death months, and computationally trivial at registry scale. Under a protocol frozen before any evaluation runs and calibrated to published national ICU registry aggregates, we compared STREAM-SMR (δ in {0.90, 0.95, 0.97}) with the risk-adjusted CUSUM and risk-adjusted EWMA across three facility-volume strata (50, 200, and 800 annual admissions) and five change scenarios (sustained steps, gradual drift, transient deterioration, and improvement). All methods were Monte-Carlo-calibrated to a common 5% false-alarm probability over a 60-month horizon, with 1,000 replications per cell.

**Results:** At δ = 0.90, STREAM-SMR matched the detection frontier of the risk-adjusted CUSUM to within one to two months across sustained-shift scenarios — median delay for an SMR step to 1.5 of 6 versus 5 months in large facilities, 14 versus 14 in medium, and 20 versus 22 in small — while returning filtered SMR estimates whose 95% credible intervals held ≥ 91% empirical coverage in every scenario–stratum cell. Empirical false-alarm probabilities were close to the 5% nominal target for all methods (range 0.035–0.066). For a three-month transient deterioration, detection was faster with STREAM-SMR conditional on occurring, but overall detection probability favored the CUSUM in large facilities.

**Conclusions:** STREAM-SMR unifies monitoring and estimation in a single Bayesian object: for a detection-delay premium of at most one to two months against the theoretically optimal CUSUM, it returns an interpretable, uncertainty-quantified SMR trajectory at every time point. The discount factor is an explicit dial between estimate smoothness and detection speed. Simulation code and the frozen protocol are publicly archived.

## 1. Introduction

National clinical registries increasingly aim to give participating facilities timely feedback on risk-adjusted outcomes rather than annual retrospective reports. For mortality, the standard quantity is the standardized mortality ratio (SMR): observed deaths divided by deaths expected under a severity-of-illness model. Cross-sectional instruments such as funnel plots [1] compare facilities at fixed reporting intervals, but by construction detect deterioration only after the reporting period closes.

Sequential alternatives adapted from industrial statistical process control address this latency [8]. The risk-adjusted CUSUM [2] accumulates patient-level log-likelihood-ratio evidence and is, for a specified step alternative, the optimal sequential detector [3]. Risk-adjusted EWMA charts [4] and the VLAD display [5] are widely used companions, including in intensive care [6].

These tools share a structural limitation: they emit an alarm, not an estimate. A CUSUM statistic of 4.2 has no direct clinical reading; after a signal, the facility must still ask *what is our SMR now, and how certain are we?* — a question the chart cannot answer without a separate estimation step. Conversely, periodically re-estimated SMRs with confidence intervals answer the estimation question but provide no principled sequential alarm.

STREAM-SMR treats the SMR as a latent state evolving in time and performs exact conjugate Bayesian filtering on it. At every month, the filtered posterior yields (i) a point estimate of the current SMR, (ii) a credible interval, and (iii) an alarm statistic — the posterior probability that the SMR exceeds a threshold — all from the same object. The method has operated in a national ICU registry environment for approximately seven years; here we characterize its operating properties in a controlled simulation against the standard sequential comparators, under a protocol frozen before any evaluation runs.

Our aim is deliberately not to show that STREAM-SMR “beats” the CUSUM at step-shift detection — optimality theory says it should not. The question is what detection-delay premium, if any, is paid for calibrated continuous estimation, and how the method’s single tuning parameter governs that trade-off.

## 2. Methods

### 2.1 Model

For a single facility, index months t = 1, …, T. Let n_t be eligible admissions, y_t observed in-hospital deaths, and E_t = Σ_i 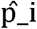 the expected deaths under a ixed severity model whose predictions 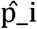 are treated as given. The estimand is the latent SMR_t, monitored on the log scale θ_t = log SMR_t.

The observation equation is y_t | λ_t ∼ Poisson(λ_t) with λ_t = E_t·exp(θ_t). State evolution follows the discount-factor construction of West and Harrison [7]: given the posterior SMR_{t−1} | D_{t−1} ∼ Gamma(α_{t−1}, β_{t−1}), the prior at t is Gamma(δα_{t−1}, δβ_{t−1}) — mean preserved, variance inflated by 1/δ — and conjugate updating gives α_t = δα_{t−1} + y_t, β_t = δβ_{t−1} + E_t.

The filtered distribution of SMR_t is therefore Gamma(α_t, β_t) in closed form, yielding the point estimate α_t/β_t, central 95% credible intervals from gamma quantiles, and the exceedance probability π_t = P(SMR_t > 1 | D_t) as the alarm statistic. A signal occurs when π_t crosses a calibrated threshold; improvement monitoring uses P(SMR_t < 1) symmetrically. Initialization was α_0_ = β_0_ = 1. Each facility-month update costs two multiplications and two additions; no linearization or sampling is required, and the update is exact for months with zero deaths — the regime in which log-transform Kalman approximations degrade and in which small facilities predominantly operate.

The discount factor δ in (0, 1] is the sole tuning parameter, with effective memory of roughly 1/(1−δ) months. We evaluated δ in {0.90, 0.95, 0.97}.

### 2.2 Comparators

All comparators consumed the same patient-level predictions 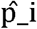. First, the risk-adjusted CUSUM [2]: a patient-sequential log-likelihood-ratio chart with odds-ratio alternative OR = 1.5 (OR = 1/1.5 for improvement monitoring). Second, a risk-adjusted EWMA in the spirit of Grigg and Spiegelhalter [4]: the monthly standardized O−E statistic smoothed with κ = 0.05. VLAD was treated as a display companion without a native alarm rule and is not tabulated; funnel-plot latency follows analytically from its annual periodicity.

### 2.3 Simulation protocol

The protocol was frozen before any evaluation runs. Three facility strata reflected the volume distribution in published national ICU registry aggregate reports: 50, 200, and 800 annual admissions, with monthly counts Poisson-distributed and case-mix risks drawn from Beta(0.5, 4.5) (mean predicted mortality 10%). True deaths were Bernoulli with probability min(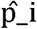·SMR_t, 0.99). Five scenarios ran over a 60-month horizon with a change point at month 25: S0, null (SMR = 1); S1a and S1b, sustained steps to 1.3 and 1.5; S2, linear drift to 1.5 over months 25–48; S3, transient elevation to 1.5 for months 25–27; and S4, an improvement step to 0.7.

#### Calibration

For each stratum and direction, every method’s threshold was set by Monte Carlo to the 95th percentile of the per-replication maximum statistic under S0 (1,000 replications) — a common in-control false-alarm probability of 5% over 60 months. This is the fairness device on which all comparisons rest.

#### Metrics

We recorded the empirical false-alarm probability (S0); the detection delay from the change point, its median, and detection probability within 6, 12, and 24 months (replications signaling before the change point were excluded and tabulated separately); and, for STREAM-SMR, the RMSE of the filtered SMR against the true path and the empirical coverage of the 95% credible interval. Each cell used 1,000 evaluation replications with fixed seeds (Python/NumPy/SciPy). Code, configuration, and the frozen protocol are archived on Zenodo (doi:10.5281/zenodo.21991702).

## 3. Results

### 3.1 Calibration held

Empirical false-alarm probabilities over 60 in-control months ranged from 0.035 to 0.066 across all fifteen method–stratum cells, against the 5% target, confirming that the Monte Carlo calibration equalized the in-control footing (Table 1).

**Table 1.**
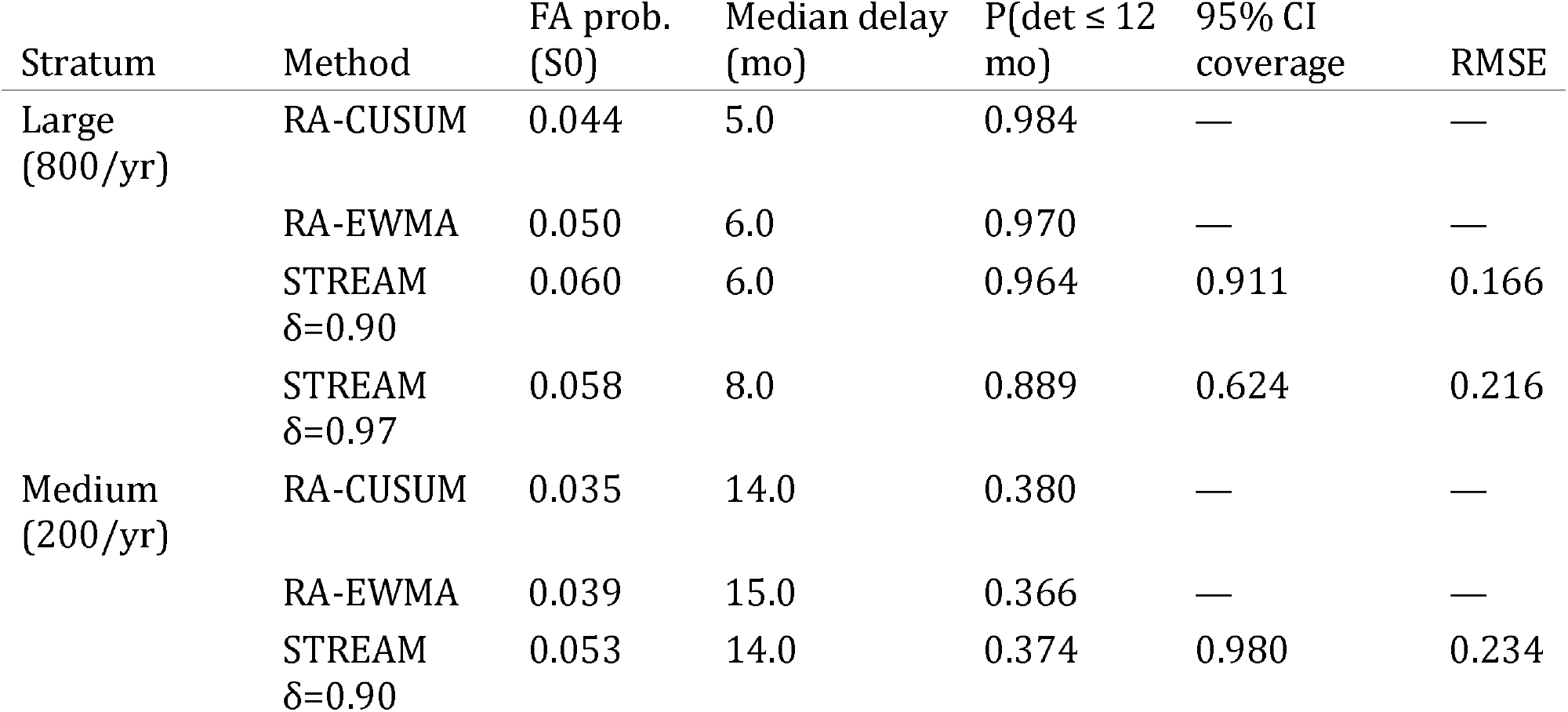
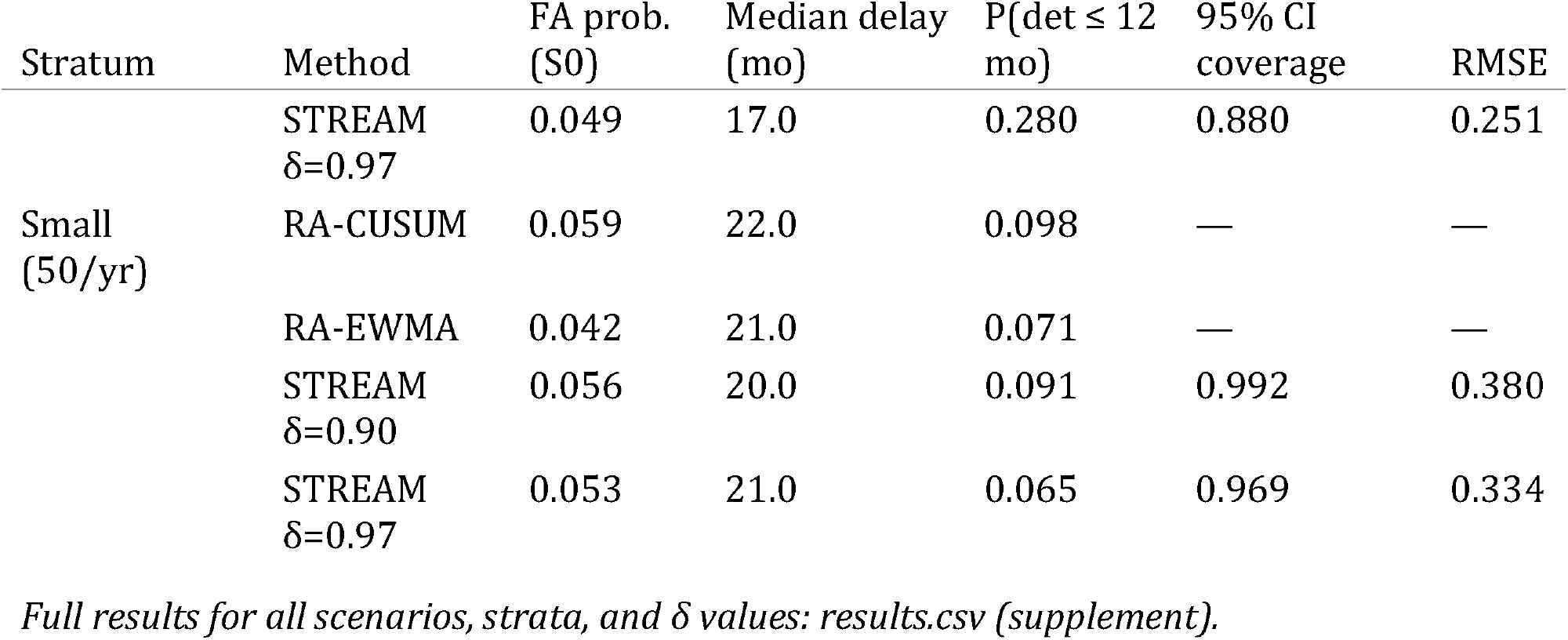
Main results (SMR 1.0 to 1.5 sustained step, S1b), by facility stratum.

### 3.2 Sustained shifts: STREAM-SMR at δ = 0.90 sits on the detection frontier

For the SMR 1.0 to 1.5 step (S1b), median detection delays were: large facilities — CUSUM 5, EWMA 6, STREAM-SMR(0.90) 6 months; medium — 14, 15, and 14; small — 22, 21, and 20. Twelve-month detection probabilities in large facilities were 0.984 (CUSUM), 0.970 (EWMA), and 0.964 (STREAM-SMR 0.90). The milder step (S1a) and gradual drift (S2) showed the same ordering with uniformly longer delays; under drift, the three methods were separated by at most two months of median delay in every stratum. Increasing δ lengthened STREAM-SMR delays monotonically — large-stratum S1b medians of 6, 7, and 8 months at δ = 0.90, 0.95, and 0.97 — the expected memory effect (Figure 3).

In the smallest stratum, STREAM-SMR(0.90) had the shortest median delay of any method under S1b (20 versus 22 months for CUSUM), though at this volume all methods detected under half of true shifts within 24 months. That floor is imposed by information content rather than method choice, and it quantifies the intrinsic limit of sequential monitoring in small facilities.

### 3.3 Transient deterioration: a genuine trade-off

For the three-month transient (S3), STREAM-SMR was faster *conditional on detection* (medium-stratum median delay 3 versus 11.5 months for CUSUM) but detected fewer episodes overall in large facilities (24-month detection probability 0.14 versus 0.25). The mechanism is structural: the CUSUM retains accumulated evidence after the episode ends, whereas the discounted filter forgets it. Neither behavior dominates; the preference depends on whether post-hoc detection of an already-resolved episode is operationally valuable.

### 3.4 Estimation: the capability the comparators do not have

At δ = 0.90, empirical coverage of the 95% credible interval was at least 0.91 in every scenario–stratum cell (range 0.911–0.996), including through shift transitions. Higher δ traded adaptivity for smoothness: at δ = 0.97, coverage during the large-stratum S1b transition fell to 0.62 because the long-memory filter lags the step, while null-scenario RMSE improved (0.087 versus 0.100 at δ = 0.90 in large facilities; 0.266 versus 0.338 in small). The CUSUM and EWMA emit no estimate and therefore have no entries in this part of the comparison — which is the substantive point (Figure 1; Table 1).

**Figure 1.**
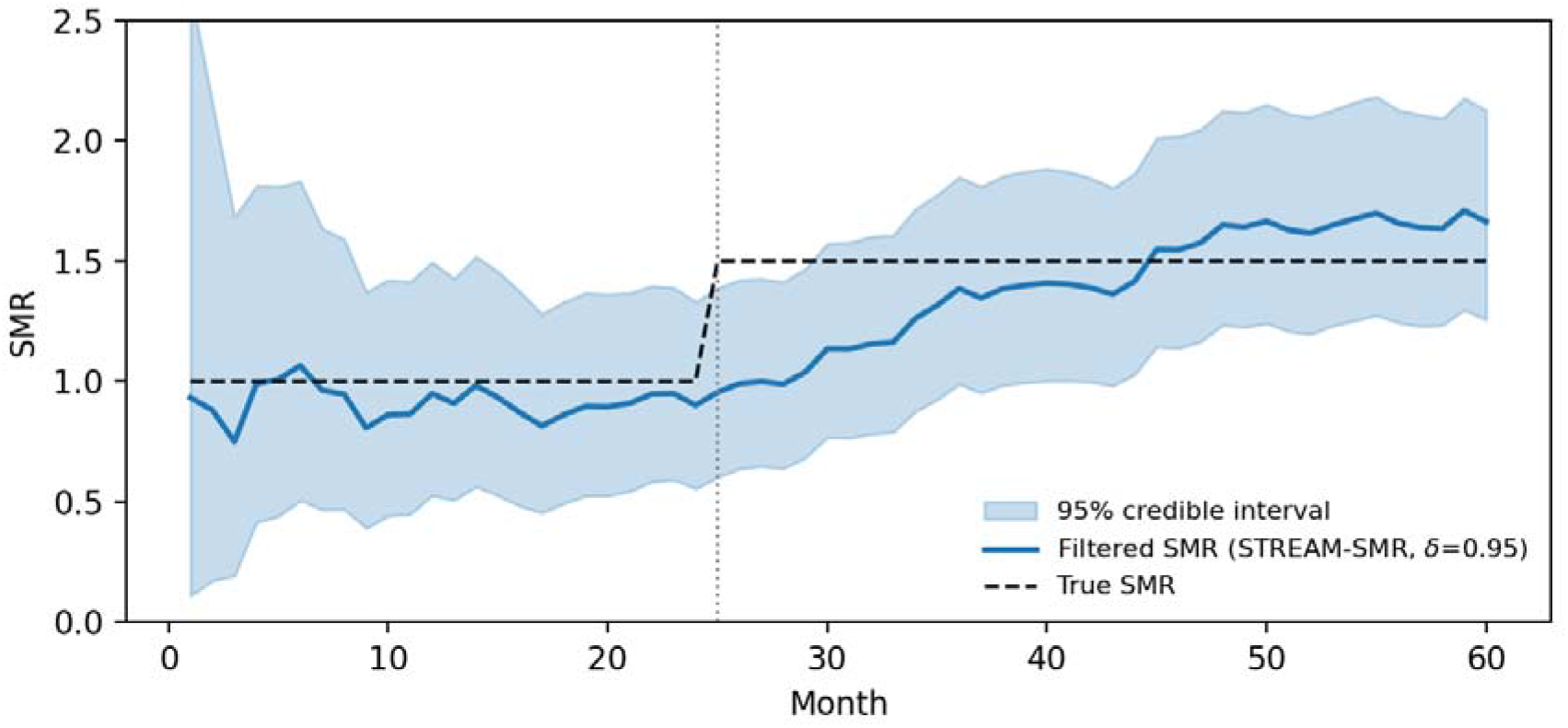
Illustrative filtered SMR trajectory with 95% credible interval (medium stratum, S1b, δ = 0.95) against the true path.

**Figure 2.**
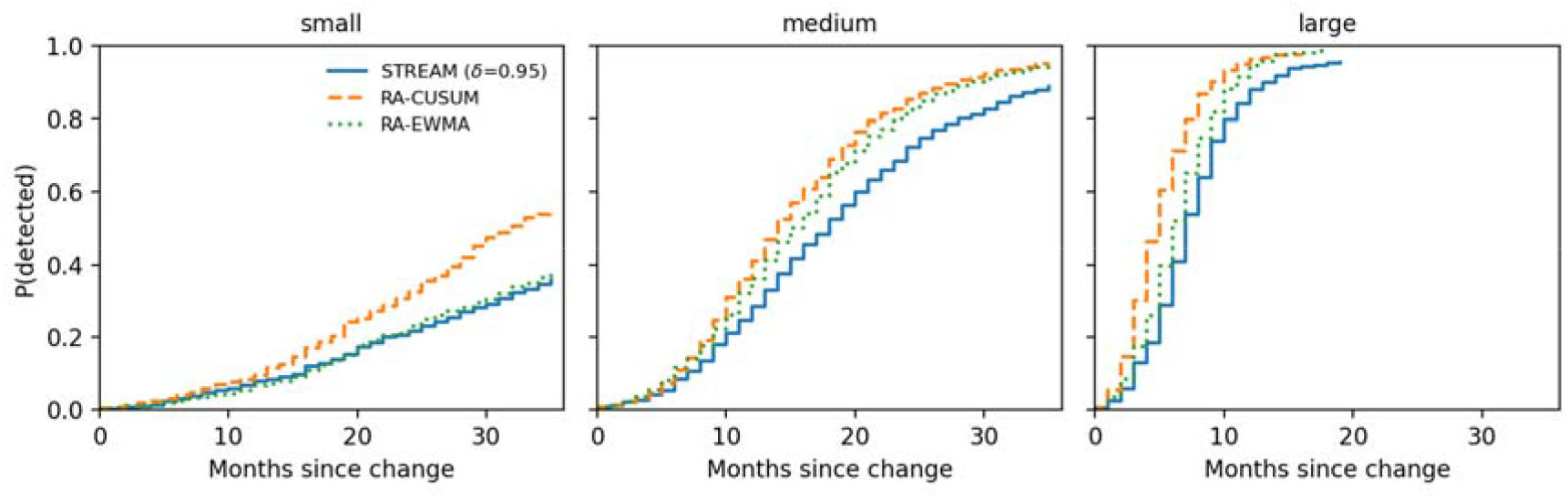
Detection-delay CDFs for the sustained 1.5 step, by stratum and method.

**Figure 3.**
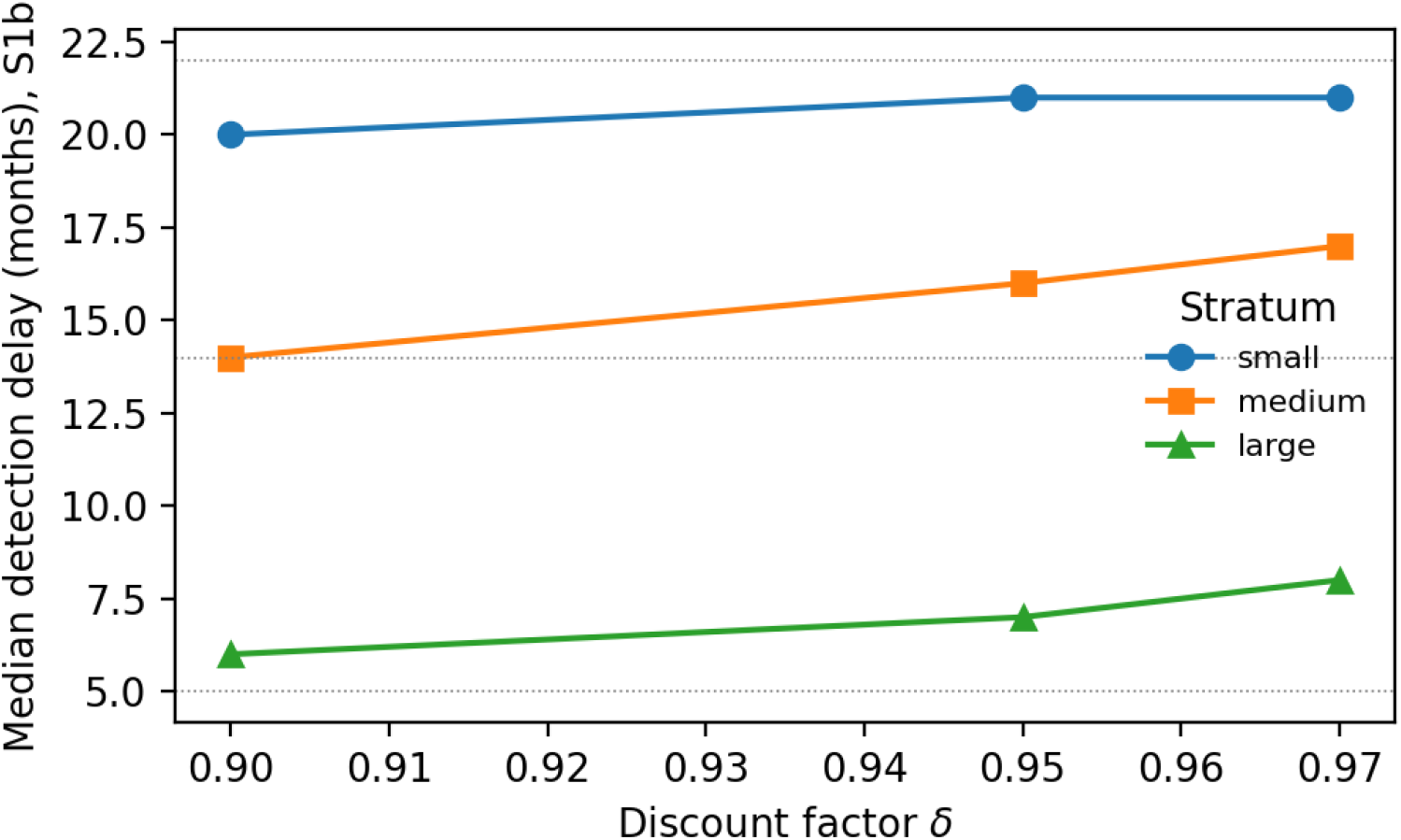
Median detection delay versus discount factor δ (S1b), with CUSUM reference lines.

## 4. Discussion

Three conclusions follow. First, the price of unified monitoring-plus-estimation is small and quantifiable: at δ = 0.90, STREAM-SMR concedes at most one to two months of median detection delay to the theoretically optimal CUSUM for sustained shifts, and concedes nothing in small facilities. Second, δ is an honest, interpretable dial — operators who prioritize alarm speed choose 0.90; those who prioritize stable published estimates choose 0.95–0.97 and accept the transition lag, with the coverage consequences quantified here. Third, calibrating the exceedance-probability alarm by in-control Monte Carlo makes it behave like a properly sized test, so the posterior probability shown to a facility and the alarm mechanism are one object rather than two disconnected artifacts.

The transient-scenario result deserves emphasis because it cuts against our method. A forgetting filter is structurally worse than an accumulating chart at flagging an already-resolved episode. In registry practice we regard this as acceptable — the estimate correctly returns to baseline, and episode review is a retrospective task — but programs whose priority is post-hoc episode capture should pair the filter with a CUSUM rather than replace it.

Several limitations bound these claims. This is a simulation study. The severity model was treated as fixed and correctly calibrated, whereas in production the risk model itself ages; miscalibration accumulating in frozen risk-adjustment infrastructure is an instance of what we have elsewhere termed institutional debt in health data systems, and disentangling model drift from true performance change is the natural next problem for state-space formulations, which can in principle carry a recalibration component in the state vector. Monthly binning, a single monitored facility (no multiplicity control across a registry’s hundreds of units), independence of admissions, and a multiplicative mortality-shift mechanism are further simplifications. Finally, the method itself has operated in a national ICU registry environment for approximately seven years; the present study characterizes the algorithm’s operating properties, not that deployment.

## 5. Conclusion

A conjugate gamma–Poisson discount filter turns SMR surveillance from alarm generation into calibrated sequential estimation, with the alarm as a by-product, at a detection cost of one to two months against the optimal step-shift detector. For registries whose obligation is to tell facilities *what their SMR is now, and with what certainty* — not merely when to worry — the trade appears worth making.

## Data and code availability

All simulation code, the frozen protocol, configuration, and seeds are openly available at Zenodo (doi:10.5281/zenodo.21991702). No patient-level data were used; facility strata were calibrated to published aggregate registry reports only.

## References

1. Spiegelhalter DJ. Funnel plots for comparing institutional performance. Stat Med. 2005;24(8):1185–1202.

2. Steiner SH, Cook RJ, Farewell VT, Treasure T. Monitoring surgical performance using risk-adjusted cumulative sum charts. Biostatistics. 2000;1(4):441–452.

3. Moustakides GV. Optimal stopping times for detecting changes in distributions. Ann Stat. 1986;14(4):1379–1387.

4. Grigg OA, Spiegelhalter DJ. A simple risk-adjusted exponentially weighted moving average. J Am Stat Assoc. 2007;102(477):140–152.

5. Lovegrove J, Valencia O, Treasure T, Sherlaw-Johnson C, Gallivan S. Monitoring the results of cardiac surgery by variable life-adjusted display. Lancet. 1997;350(9085):1128–1130.

6. Cook DA, Steiner SH, Farewell VT, Morton AP. Monitoring the evolutionary process of quality: risk-adjusted charting to track outcomes in intensive care. Crit Care Med. 2003;31(6):1676–1682.

7. West M, Harrison J. Bayesian Forecasting and Dynamic Models. 2nd ed. New York: Springer; 1997.

8. Grigg O, Farewell VT, Spiegelhalter DJ. Use of risk-adjusted CUSUM and RSPRT charts for monitoring in medical contexts. Stat Methods Med Res. 2003;12(2):147–170.

